# Factors associated with persistent hypertension one year postpartum in persons with gestational hypertension or preeclampsia

**DOI:** 10.1101/2022.10.06.22280786

**Authors:** M Christine Livergood, Leigh Mahlum, Jordan Hauck, Maggie Tallmadge, Kara Hoppe, Anna Palatnik

**Affiliations:** Department of Obstetrics and Gynecology, Medical College of Wisconsin, Milwaukee, Wisconsin; Division of Maternal-Fetal Medicine, Department of Obstetrics and Gynecology, University of Wisconsin Madison, Madison, Wisconsin

**Keywords:** gestational hypertension, preeclampsia, postpartum, persistent hypertension, stage I, hypertension, antihypertensive medications, hypertensive disorders of pregnancy

## Abstract

**Objective:** To identify factors associated with persistent hypertension one-year postpartum after pregnancy complicated by gestational hypertension or preeclampsia.

**Study Design:** A retrospective case-control study of postpartum patients who had a diagnosis of gestation hypertension or preeclampsia during a recent pregnancy and attended one-year postpartum annual exam and blood pressure check between 2014 and 2019 in a single academic medical center. Cases were defined as persons with persistent hypertension one year postpartum, using the 2017 American College of Cardiology/American Heart Association (ACC/AHA) guidelines, defining stage I hypertension as systolic blood pressure ≥130 mmHg or diastolic blood pressure ≥80 mmHg. Controls were defined as non-pregnant persons with normal blood pressure (BP) at one year postpartum. Using bivariate and multivariate analyses, demographic, clinical and labor characteristics were compared between persons who had persistent hypertension one-year postpartum and controls.

**Results:** Of the 595 persons included in this analysis, 268 (45.0%) had persistent hypertension one year postpartum. Bivariate analyses demonstrated that older maternal age, higher body mass index (BMI) at first prenatal visit, at delivery, and one year postpartum, mild-range BP (compared to normal BP) prior to discharge, and patients with elevated BP at 6-week postpartum visit, were more likely to have persistent hypertension one-year postpartum. In contrast, nulliparity was associated with lower risk of having persistent hypertension at one-year postpartum. Multivariate logistic regression demonstrated that mild range BP prior to discharge (aOR 1.78, 95%CI 1.16-2.72), elevated BPs at 6 weeks postpartum (aOR 2.01, 95% CI 1.36-3.00), and higher BMI at one-year postpartum (aOR 1.07, 95%CI 1.00-1.14), remained to be significantly associated with higher odds of persistent hypertension one-year postpartum, while nulliparity remained to be associated with lower odds of persistent hypertension one-year postpartum (aOR 0.55, 95%CI 0.36-0.84)

**Conclusion:** In this cohort, 45% of patients with gestational hypertension or preeclampsia had persistent hypertension one-year postpartum by the 2017 ACC/AHA hypertension definition. Patients that had mildly elevated BPs in the immediate postpartum period as well as at 6 weeks postpartum, and higher BMI one year postpartum, had higher risk of having at least stage I HTN one year following pregnancy complicated by gestational hypertension or preeclampsia.

## Introduction

Hypertensive disorders of pregnancy (HDP), referred in this paper to gestational hypertension and preeclampsia, are one of the leading causes of maternal and perinatal mortality worldwide.^1,2^ In the United States, HDP complicate 9.12% of all pregnancies, but accounts for 16% of all maternal deaths.^1,2^ Between 1987 and 2004, the rate of preeclampsia increased by 25%, and in comparison with pregnant people giving birth in 1980, those giving birth in 2003 were 6.7 times more likely to be diagnosed with preeclampsia with severe features.^2^ The increasing incidence of preeclampsia stresses the importance of understanding the impact of HDP on both the women’s cardiovascular health as well as the burden to the healthcare system.

The effects of HDP are not limited to pregnancy and are associated with a lifetime increased incidence of chronic hypertension and cardiovascular disease, including coronary heart disease and stroke.^2^ Prior research has demonstrated that a substantial proportion of persons with preeclampsia will have persistent hypertension up to five years after delivery and long-term risk of developing chronic hypertension.^3,4^ In these studies, associated risk factors for persistent hypertension included excessive gestational weight gain, presence of gestational diabetes mellitus, preeclampsia with severe features, dyslipidemia and age greater than 40.^4-6^ Identifying modifiable risk factors for persistent hypertension may help to reduce long-term consequences and the associated health burden of postpartum preeclampsia.

In 2017, the American College of Cardiology/American Heart Association (ACC/AHA) updated their guidelines for the prevention, detection, evaluation, and management of high blood pressure in adults.^7^ These guidelines defined normal blood pressure as <130/80 mmHg and lowered the threshold for a diagnosis of chronic hypertension to systolic blood pressure (SBP) 130-139 mmHg or diastolic blood pressure (DBP) 80-89 mmHg (stage 1).^7^ The prior qualifying diagnosis of hypertension; SBP ≥140 mmHg or DBP ≥90 mmHg is now classified as stage 2 hypertension.^7^ Despite the ACC/AHA guideline changes, classification of HDP still uses SBP≥140 or DBP ≥90 to confirm the diagnosis of hypertension. Since most studies looking at the risk of chronic hypertension following HDP used the older and higher definition of stage I hypertension (≥140/90 mmHg), the objective of this study was to determine the rate of persistent/chronic hypertension one-year postpartum following HDP and identify risk factors associated with persistent hypertension, using the 2017 ACC/AHA guidelines for hypertension definition.

## Methods

This was a retrospective case-control study of postpartum persons diagnosed with HDP during pregnancy, intrapartum, or postpartum, between January 2014 and December 2019 at a single Midwest academic medical center. Cases were defined as persons who remained hypertensive (stage 1 or higher) at 1-year postpartum by the ACC/AHA guidelines.^7^ Controls were defined as persons with normal blood pressures, defined as <130 mmHg SBP and <80 mmHg DBP at 1-year postpartum. Institutional review board approval was obtained prior to the initiation of the study.

Persons were included in the analysis if they were 18 years or older, received prenatal care at our academic institution or one of the affiliated community clinics and were diagnosed with HDP during pregnancy, intrapartum, or during their postpartum hospital stay, using ICD9 and ICD10 codes for diagnosis of HDP. Persons were excluded if they had chronic pregestational hypertension, lack of follow-up one year postpartum, or were pregnant at one year postpartum (Figure 1).

**Figure 1.**
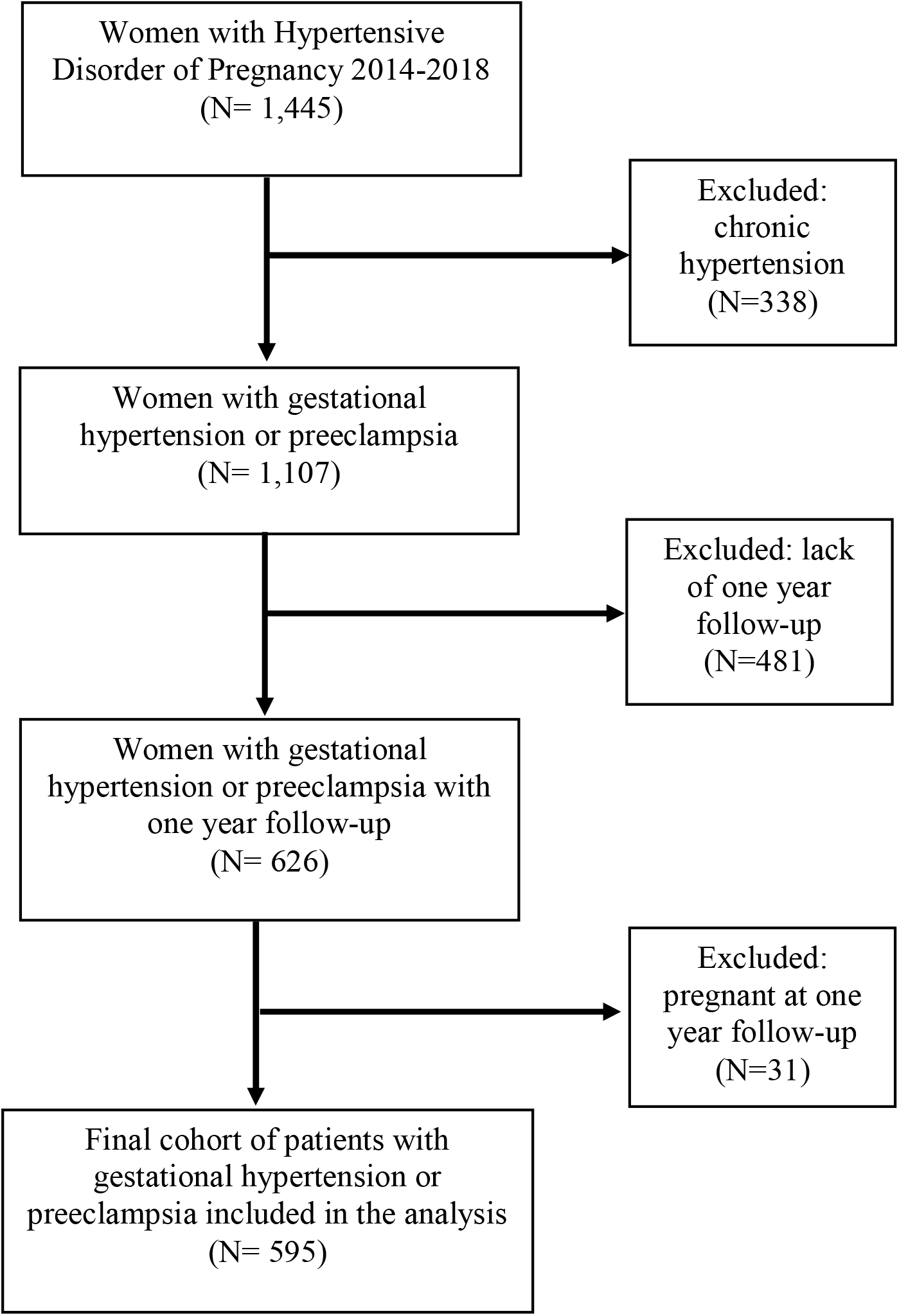
Flowchart of persons included in the study

Persons’ demographics, clinical characteristics and delivery outcomes were abstracted from the electronic medical records. The demographics included the maternal age, maternal race and ethnicity, marital status, and insurance type. Clinical characteristics and delivery outcomes included gestational age at delivery, parity, early pregnancy BMI obtained at the first prenatal visit, BMI at delivery, and BMI one-year postpartum, type of HDP diagnosis (with or without severe features), magnesium sulfate administration during hospital admission, initiation of antihypertensive pharmacotherapy after delivery and prior to discharge, and blood pressures data during delivery admission and at 6-weeks postpartum. Mild-range blood pressure prior to discharge after delivery hospitalization was defined as blood pressure in the range of 140s-150s mmHg SBP or 90s-100s mmHg DBP. This was chosen as a variable since there are currently no guidelines that these blood pressures need to be treated postpartum and patient may be discharged home without antihypertensive medications for mild-range blood pressures.^2^ In addition, we collected how many persons were taking antihypertensive medications one-year postpartum.

All analyses were performed with Stata version 14.0 (StataCorp College Station, TX). Cases and controls were compared in bivariate analysis using Student t test, χ^2^ test, Fischer exact test and Wilcoxon rank sum (Mann-Whitney U) when appropriate. Bivariate and multivariate logistic regression was performed and unadjusted and adjusted odds rations with 95% confidence intervals were reported. Variables were included in the multivariate logistic regression model if they were clinically associated with the chance of persistent hypertension or if they had p<0.05 in the bivariate analysis. All tests were 2-tailed and a p<0.05 was used to define significance.

## Results

During the study period, approximately 12,500 deliveries occurred in our institution with a total of 1,445 persons with HDP or chronic hypertension during pregnancy (11.6%). After excluding persons with chronic hypertension (n=338), persons that were lost to follow-up one year postpartum (n=481), and persons who became pregnant at one year postpartum (n=31), a total of 595 persons were eligible for analysis (Figure 1). Of these, 268 (45.0%) had persistently elevated blood pressure at 1-year postpartum using the ACC/AHA 2017 guidelines. Table 1 describes maternal characteristics stratified by elevated blood pressures at 1-year postpartum. Persons with elevated blood pressure at 1-year postpartum were older, had higher BMI at first prenatal visit, at delivery, and at one year postpartum, and less likely to be nulliparous. In addition, persons with persistent hypertension were more likely to be discharged with mild-range blood pressures (58.9% vs. 44.9%, p<0.001) during their delivery hospitalization, in the first 2-4 days postpartum. They also continued to have higher blood pressures at 6 weeks postpartum. A total of 64 (10.8%) persons of all persons included in this analysis were taking antihypertensive medications at one-year postpartum, with higher percent in the group of elevated blood pressures (Table 1).

**Table 1:** Bivariate analysis for the outcome of persistent hypertension at 1-year postpartum in patients with gestational hypertension or preeclampsia.

| | Elevated BP 1-year postpartum<br>($\geq 130/80$ mmHg)<br>(N=268) | Normal BP 1-year postpartum<br>(N=327) | P value |
| --- | --- | --- | --- |
| Maternal age (y) | **31.3 ± 5.4** | **29.4 ± 5.7** | **< 0.001** |
| BMI at first prenatal visit (kg/m<sup>2</sup>) | **31.8 ± 7.4** | **29.4 ± 7.5** | **< 0.001** |
| BMI at delivery (kg/m<sup>2</sup>) | **35.9 ± 7.0** | **34.0 ± 7.6** | **< 0.001** |
| Race |  |  | 0.211 |
| Non-Hispanic White | 163 (61.1) | 225 (68.8) |  |
| Non-Hispanic Black | 80 (30.0) | 78 (23.9) |  |
| Hispanic | 3 (1.1) | 5 (1.5) |  |
| Other | 21 (7.8) | 19 (5.8) |  |
| Gestational age at delivery (wks) | 37.6 (36.1, 38.9) | 37.9 (36.3, 39.1) | 0.102 |
| Nulliparity | **114 (42.5)** | **206 (63.0)** | **< 0.001** |
| Insurance |  |  | 0.391 |
| Public | 172 (64.2) | 221 (67.6) |  |
| Private | 95 (35.4) | 106 (32.4) |  |
| None | 1 (0.4) | 0 (0.0) |  |
| Preeclampsia with severe features, HELLP or eclampsia | **167 (62.3)** | **101 (37.7)** | **0.774** |
| Magnesium sulfate given intrapartum or postpartum | 112 (41.8) | 130 (39.8) | 0.469 |
| Mild range BP on day of discharge (compared to normal range) | **158 (58.9)** | **146 (44.9)** | **0.001** |
| Discharged home on antihypertensive medications | 57 (21.6) | 51(15.7) | 0.066 |
| Elevated BP $\geq 130/80$ mmHg at 6 weeks postpartum | **143 (60.6)** | **102 (36.0)** | **<0.001** |
| BMI at one year postpartum (kg/m<sup>2</sup>) | **32.6 ± 7.9** | **29.6 ± 7.8** | **<0.001** |
| Taking antihypertensive medications 1 year postpartum | **43 (16.4)** | **21 (6.6)** | **<0.001** |

All data presented as N (%), mean ± SD, or median (IQR)
BMI= body mass index, HELLP = hemolysis, elevated liver enzymes and low platelets syndrome, BP = blood pressure; bold represents statistical significance

Table 2 describes unadjusted and adjusted odds ratios for the risk factors of persistent hypertension one-year postpartum. In bivariate analysis, older age, higher BMI early in pregnancy, at delivery, and at one-year postpartum, multiparity, mild-range blood pressures prior to discharge from delivery hospitalization, as well as at 6-weeks postpartum visit, were significantly associated with persistent hypertension one-year postpartum. In the adjusted analysis, after controlling for confounding factors, including the factor of taking antihypertensive medications one-year postpartum, mild-range blood pressures prior to discharge (aOR1.78, 95%CI 1.16-2.72), and at 6 weeks postpartum (aOR 2.01, 95%CI 1.36-3.00) remained to be significantly associated with persistent hypertension one year postpartum. In addition, higher BMI one year postpartum was also associated with persistent hypertension one-year postpartum (aOR 1.07, 95%CI 1.00-1.14). Nulliparity remained to be associated with lower odds of persistent hypertension one-year postpartum (aOR 0.55, 95% CI 0.36-0.84).

**Table 2:** Unadjusted and adjusted logistic regressions of factors associated with persistent hypertension at 1-year postpartum

|  | Unadjusted Odds Ratio<br>(95% CI) | Adjusted Odds Ratio<br>(95% CI) |
| --- | --- | --- |
| Maternal age | 1.06 (1.03 – 1.09) | 1.02 (0.98 – 1.06) |
| BMI early pregnancy | 1.05 (1.03 – 1.06) | – |
| BMI at delivery | 1.04 (1.02 – 1.06) | 0.97 (0.91 – 1.04) |
| Race |  |  |
| Non-Hispanic White | Ref | Ref |
| Non-Hispanic Black | 1.41 (0.97 – 2.05) | 1.27 (0.77 – 2.12) |
| Hispanic | 0.83 (0.20 – 3.51) | 0.30 (0.05 – 1.79) |
| Other | 1.52 (0.80 – 2.93) | 1.13 (0.51– 2.47) |
| Gestational age at delivery | 0.97 (0.92 – 1.01) | 0.97 (0.89 – 1.06) |
| Nulliparity | **0.43 (0.31 – 0.60)** | **0.55 (0.36 – 0.84)** |
| Insurance |  |  |
| Private | Ref | – |
| Public | 1.15 (0.82 – 1.62) |  |
| None | NA |  |
| Preeclampsia with severe features, HELLP or eclampsia | 0.95 (0.68 – 1.32) | 0.44 (0.15 – 1.33) |
| Magnesium sulfate given intrapartum or postpartum | 1.16 (0.87 – 1.55) | 1.91 (0.64 – 5.67) |
| Mild range BPs on day of discharge (compared to normal range) | **1.76 (1.27 – 2.44)** | **1.78 (1.16 – 2.72)** |
| Discharged on antihypertensive medications | 1.48 (0.97 – 2.25) | 0.91 (0.53 – 1.58) |
| Elevated BP ≥ 130/80 mmHg at 6 weeks postpartum | **2.73 (1.91 – 3.90)** | **2.01 (1.36 – 3.00)** |
| BMI 1 year postpartum | **1.04 (1.03 – 1.07)** | **1.07 (1.00 – 1.14)** |
| Taking antihypertensive medications 1 year postpartum | **2.78 (1.61 – 4.82)** | 1.85 (0.90 – 3.80) |

HELLP= hemolysis, elevated liver enzymes and low platelets syndrome, BP= blood pressure; bold represents statistical significance

## Discussion

In this study, we found that 45.0% of persons with HDP have persistent hypertension one-year postpartum using the 2017 ACC/AHA guidelines. We also found that mild-range blood pressures prior to discharge after delivery hospitalization (approximately postpartum days 2-4), elevated blood pressures (≥130/80 mmHg) at 6 weeks postpartum, and higher BMI one-year postpartum were significantly associated with persistent hypertension one-year postpartum.

Our study joins a growing body of literature examining factors associated with postpartum hypertension following HDP.^8-16^ Goel and colleagues demonstrated that 50% of persons with hypertension of any etiology in pregnancy remained hypertensive at 6 weeks postpartum.^9^ The risks for persistent hypertension in this cohort were higher BMI, history of hypertension and history of diabetes.^9^ Benschop and colleagues demonstrated that there was a 41.5% rate of persistent hypertension 1-year following a pregnancy complicated by preeclampsia with severe features.^10^ Black and colleagues followed persons with and without HDP and demonstrated that patients with HDP were 2.36 more likely to develop hypertension in the year following delivery.^11^ In a recent review article examining HDP and future cardiovascular risk, there were several factors that were identified to be associated with development of chronic hypertension, including early-onset preeclampsia compared to late onset preeclampsia and recurrent preeclampsia compared to persons who had a successive unaffected pregnancy.^12-15^ As there is some debate as whether baseline characteristics that put one at risk for HDP are also predisposing the person to further cardiovascular risk, Mito and colleagues found that persons with HDP had a higher risk of developing hypertension at 5-years even when adjusting for age, BMI, family history of hypertension and salt intake.^3^ Similarly, Bergen and colleagues found that adjusting for BMI attenuates the relationship between HDP and subsequent diagnosis of chronic hypertension.^16^

Interestingly, our analysis indicates preeclampsia with severe features was not associated with persistent hypertension at one-year postpartum, while presence of mild-range blood pressures (140s-150s mmHg SBP or 90s-100s mmHg DBP) prior to hospital discharge after delivery was associated with higher odds of persistent hypertension, independent of type of HDP with which the patients were diagnosed. We hypothesize that this can be explained by the fact that preeclampsia with severe features was most likely associated with severe-range BP and led to initiation of postpartum antihypertensive medications and perhaps closer postpartum follow-up. In contrast, postpartum patients with mild-range blood pressures prior to discharge were not started on antihypertensive medications and this could have contributed to slower recovery and long-lasting endothelial dysfunction postpartum secondary to HDP. Interestingly, even after adjusting for intake of antihypertensive medications, mild-range blood pressures prior to discharge remained to be associated with persistent hypertension one-year postpartum.

Our study also shows that an elevated blood pressure at the 6-week postpartum visit is a risk for persistent hypertension as well as higher BMI one year postpartum. Both of these risk factors are modifiable. Trials are needed to investigate whether medical treatment of postpartum hypertension potentially to achieve a BP that is less than stage 1 AHA/ACC BPs in the first year or intensive lifestyle modification and weight loss in the first year postpartum can reverse or reduce cardiovascular sequala of HDP.

Our study has several strengths. First, we analyzed our data with the focus on newer diagnosis of stage I hypertension using the 2017 ACC/AHA guidelines. Second, our sample size was large and racially diverse with detailed data on blood pressure values during the delivery related hospital admission, 6-weeks postpartum, and one-year postpartum. Third, through detailed chart review we were able to use exact diagnoses of HDP types in our analysis, differentiating preeclampsia with and without severe features. Fourth, we controlled for intake of antihypertensive medications and BMI at one-year postpartum. However, we also had a few limitations. First, we had a very small representation of Hispanic persons in our cohort. Second, a portion of patients was lost to follow-up or became pregnant one-year postpartum (Figure 1), and we could not report their hypertension status at one-year postpartum. Third, this study was conducted in a retrospective fashion and carries with it the limitations of this study design including inability to form causal associations.

In conclusion, we identified several modifiable risk factors for persistent hypertension one-year postpartum following HDP. While HDP is a known risk factor for future cardiovascular events, identifying persons who will have persistent hypertension following their pregnancy and optimizing their management in the first year postpartum period may be an avenue to attempt to mitigate the long-term sequalae of HDP.

## Data Availability

All data produced in the present work are contained in the manuscript

## Notes

The authors did not report any potential conflict of interest.

### Competing Interest Statement

The authors have declared no competing interest.

### Funding Statement

This study did not receive any funding

### Author Declarations

Ethics committee/IRB of Medical College of Wisconsin gave ethical approval for this work.

